# Use of Implantable Loop Recorders: Incidence, Indications for Use, and Pre-implant Rhythm Monitoring Across U.S. Health Systems from 2020-2025

**DOI:** 10.64898/2026.09.25.26364058

**Authors:** Matthew C. Andersen, Joseph S. Ross, Rohini Ghosh, Luis Correia, Sanket S. Dhruva

**Author notes:** **Contact information for corresponding author:**Sanket Dhruva, MD, MHS, 4150 Clement St, 111C, San Francisco, CA 94121.

## Abstract

**Background:** Implantable loop recorders (ILRs) provide continuous rhythm monitoring. Professional society guidelines and expert consensus statements recommend ILR use for patients with recurrent syncope and cryptogenic stroke, but recommendations are weaker for other indications. We assessed ILR use over time and concordance between indication and clinical guidance.

**Methods:** We conducted a retrospective cohort study using Epic Cosmos data from 1/1/2020-12/31/2025. Among active patients with at least one of 13 diagnoses potentially eligible for ILR placement, we identified ILR placements and classified them by concordance with professional society guidance: concordant, possibly, or non-addressed. Annual incidence was estimated overall and by indication. We also described the proportion of patients who received longer-term (>7 days) rhythm monitoring in the year prior to placement.

**Results:** Of 47,046,706 patients with diagnoses potentially eligible for ILR placement, 201,103 (0.4%) had an ILR placed. ILR placements increased from 3.08 ILRs per 100,000 patient-days in 2020 to 4.13 in 2025 (5.9% annual growth). The most common indications were cryptogenic stroke (28.6%) and atrial fibrillation without prior ablation (21.4%). Overall, 43.9% were guidance concordant, 28.5% possibly concordant, and 24.8% non-addressed. Relative annual growth in ILR placements per 100,000 patient-days was significant across all guidance categories: concordant (1.3%, 95% CI, 0.9%-1.7%), possibly concordant (5.5%, 95% CI, 5.0%-6.0%) and non-addressed (7.4%, 95% CI, 6.8%-8.0%). Among 125,631 outpatient placements, 50,181 (39.9%) had any prior long-term rhythm monitoring.

**Conclusion:** ILR placements are uncommon but increasing, with faster annual growth for indications that may not be guidance concordant. More than half of patients lacked long-term rhythm monitoring prior to outpatient ILR placement.

**What is known:**

– Prior research through 2018 demonstrated that many ILRs were placed for indications not supported by professional society guidelines or expert consensus statements.
– Implantable loop recorder (ILR) technology has advanced with longer battery life, easier implantation processes, and robust remote monitoring and reprogramming.

**What the Study Adds:**

– ILR use remains uncommon for patients with diagnoses potentially eligible for ILR placement.
– Annual incidence of ILR placement is increasing, especially among patients with indications not addressed by professional society guidelines or expert consensus statements.
– Most ILR placements occur without pre-implant rhythm monitoring.

## Background

Implantable loop recorders (ILRs), also known as insertable cardiac monitors, are small subcutaneously implanted devices that continuously monitor patients’ heart rhythms for up to 6 years. These devices were first approved by the Food and Drug Administration (FDA) in 1998 **(Supplemental Table 1)**. Over time, they have gained extended battery longevity, utilized improving diagnostic algorithms to reduce false alerts,^1–3^ and added remote reprogramming capabilities to optimize alerts.^4,5^ Concurrently, ILRs have become smaller and implantation procedures shorter, with minimal complication risk.^6^ In 2025, the mean Medicare reimbursement for placement and programming ranged from $4,016 in a physician’s office to $8,455 in a hospital outpatient department.^7^ The 2025 ILR market size was estimated at $1.9 billion, and expected to nearly double by 2030.^8^

Multiple professional specialties provide guidance about which clinical indications are best suited for monitoring using ILRs, including Class I recommendations in evaluating cryptogenic stroke, recurrent syncope, or syncope with high risk criteria.^9–14^ Recommendations for other indications vary, but there is lower strength of evidence for unexplained falls, palpitations, supraventricular tachycardia, and surveillance of atrial fibrillation following ablation.^9,11,15–18^ A prior study found that approximately one-fourth of ILRs placed from 2011 through 2018 were for clinical indications not addressed by professional society guidelines.^19^ Despite continued advances in ILR technology and ease of placement,^1–6^ there are no contemporary data about practice patterns.

Accordingly, we sought to characterize trends in ILR placement over the past 6 years within a national population, examining overall placement incidence, including by indication and setting. Among patients receiving ILRs, we stratified placement by concordance with professional society guidelines and expert consensus statements. Further, given advances in non-invasive rhythm monitoring,^20^ we examined what monitoring that patients had received prior to ILR placement.

## Methods

### Data Source

We used January 1, 2020 through December 31, 2025 data from Epic Cosmos, representing a community of health systems with Epic electronic health records, which dynamically updates as health systems’ data are continually added, deduplicating patient records across multiple health systems.^21–23^ Data were retrieved on March 28, 2026. Epic Cosmos requires participating institutions to submit a consistent span of recent historical data, while allowing—but not mandating—the backloading of older records.^21^ As of December 2025, Epic Cosmos included data from more than 300 million patients who received care at more than 2,000 hospitals and 52,000 clinics, of which >99% are located in the U.S. The study was reported in accordance with the RECORD statement and was exempt from ethics review and informed consent because it used nonidentifiable data.

### Cohort Construction

We first identified all patients who had received any of 13 diagnoses for a clinical indication potentially eligible for ILR placement (**Table 1**); we excluded patients younger than 18 years, who were missing age or sex data, or who had placement of pacemaker or implantable cardioverter-defibrillator prior to eligible diagnosis (**Supplemental Table 2**). Clinical indications were determined based on prior research^19^ and professional society guidelines and expert consensus statements:^9–14,16–18^ cryptogenic stroke, recurrent syncope, syncope, stroke or transient ischemic attack (TIA), non-sustained ventricular tachycardia (VT) or frequent premature ventricular contractions (PVCs), palpitations, uncontrolled epilepsy, recurrent falls, dizziness, atrial flutter, supraventricular tachycardia, atrial fibrillation (AF) prior to ablation and AF with ablation (**Supplemental Table 3**). Potentially eligible clinical indications were determined from billed International Classification of Diseases (ICD)-10 diagnosis codes for any clinician encounter, including hospital, emergency department, and ambulatory settings.

**Table 1.**
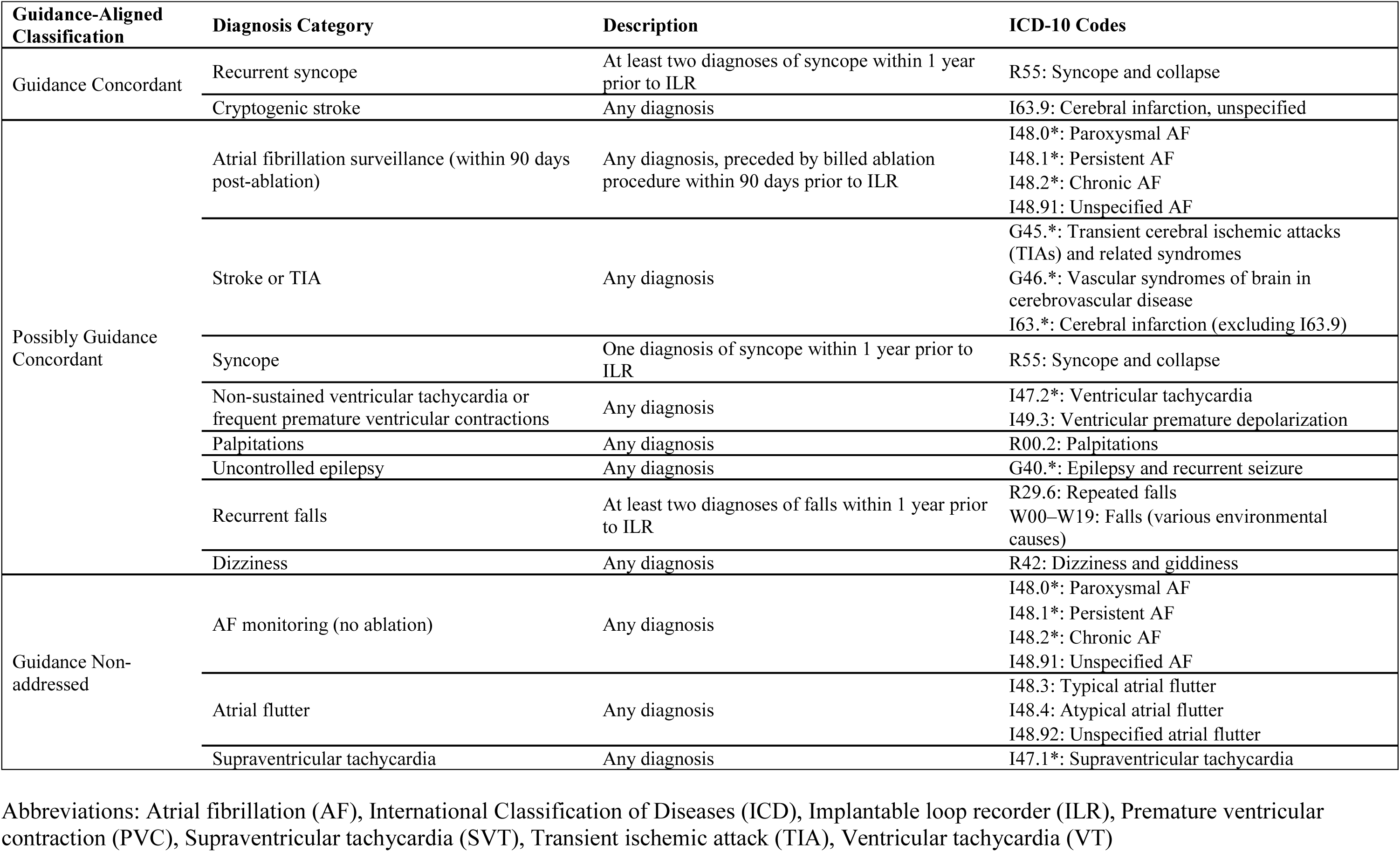
Implantable Loop Recorder-Eligible Diagnosis Categories and Guidance-Concordance Classifications.

Next, we identified all patients who underwent ILR placement based on procedure codes. Among these patients, we then restricted to those who were active in the health system, defined as any clinical encounter with an ICD-10 diagnosis code within the 1-year period preceding (as early as January 1, 2019) and following ILR placement as documented in Epic Cosmos, not including the encounter with ILR placement. ILR placement for AF within 90 days of ablation was classified as ‘AF post-ablation surveillance’ and was considered a distinct indication from placement for AF without near-term ablation (which was classified as ‘AF monitoring’). For patients with multiple ILR placements, only the first placement was eligible for inclusion.

### Outcomes

#### Denominator of Patients Eligible for ILR Placement

We calculated incidence of ILR placement among all patients who had received any of 13 diagnoses for a clinical indication potentially eligible for ILR placement (as described above), overall and stratified by clinical indication and setting. All patients with any qualifying clinical indication contributed at-risk patient time to the denominator, regardless of whether they ultimately underwent ILR placement. Patients entered the at-risk cohort on the date of their first qualifying clinical indication diagnosis and accrued a maximum of 90 days of person-time from that date. This method assumes ILR placement within one annual quarter after a qualifying diagnosis, consistent with prior research.^19^ For ILRs placed following recurrence of a qualifying clinical indication (i.e., recurrent syncope and recurrent falls), we identified the initial occurrence with no preceding diagnosis within a 1-year look-back period. We considered the first subsequent diagnosis within 1 year of the initial event to be a recurrent event, and that recurrent event contributed 90 days of person-time to the at-risk pool. We excluded ILR without any of the 13 diagnoses from calculations of incidence.

We censored patients at the earliest date of ILR placement; pacemaker or implantable cardioverter-defibrillator placement, given that these devices monitor cardiac rhythm; or documented death.

#### Indication for ILR Placement

Among patients who underwent ILR placement, we determined the clinical indication for placement. For outpatient ILR placements, patients were required to receive any qualifying diagnosis within the same diagnosis category at ≥2 different times; clinical indication was determined based on diagnostic coding during the encounter and within the 90 days preceding ILR placement. For the two indications that required event recurrence (i.e., recurrent syncope and recurrent falls), patients were required to receive the qualifying diagnosis code at ≥3 different times: ≥2 different times in the preceding 1 year, in addition to at ILR placement. For inpatient ILR placements, clinical indication was determined based on diagnostic coding during the encounter.

Patients who did not meet these criteria were categorized as “Non-Classifiable”. If patients received diagnoses for multiple qualifying clinical indications, they were assigned based on the diagnosis for which clinical practice guidelines or expert consensus more strongly recommended ILR placement (e.g. cryptogenic stroke rather than dizziness). If recommendations were consistent for multiple clinical indications, we assigned the diagnosis with greater clinical severity (e.g. TIA rather than palpitations).

#### Guidance Concordance for ILR Placement

Patient clinical indications for ILR placement were classified by concordance based on clinical practice guidelines and expert consensus statements using three categories based on a previously published classification scheme:^19^ 1) concordant, 2) possibly concordant, or 3) non-addressed **(Table 1)**. We determined guidance concordance by first reviewing the most recent U.S. and European cardiology professional society guidelines (**Supplemental Table 3**).^9–14,16–18^

We classified ILR placement as *concordant* when the indication was supported by at least one class I (“is recommended”) or IIa (“is reasonable”) recommendation. We classified ILR placement as *possibly concordant* when the diagnosis carried a class IIb (“may be reasonable”) recommendation. In the absence of society guidelines, we relied on expert consensus statements, classifying “should do” recommendations as *concordant* and “should consider” recommendations as *possibly concordant*. We considered ILR placements without society guideline or expert consensus recommendations for ILR use as *guidance non-addressed*. ILR with non-classifiable diagnoses were not assigned a guidance concordance classification.

#### Pre-ILR Monitoring and Post-ILR Placement Follow-Up

For all patients who received ILR placement, we assessed rhythm monitoring within one year prior to the procedure, classified as short-term (≤2 days), medium-term (>2 and ≤ 7 days), or long-term (>7 days). To ensure sufficient data prior to ILR placement, we limited this analysis to patients who were active in the health care system at least one year prior to placement. Patients with multiple monitoring modalities were assigned the longest-duration category.

In addition, in the 2 years following ILR placement we determined rates of follow-up events, including interrogation, reprogramming, and removal. To obtain the most accurate estimates, we limited these analyses to patients who were actively followed by cardiologists, determined in Epic Cosmos by visits at least 2 years following ILR placement in a department categorized as cardiology or clinician in a cardiology sub-specialty.

Rates of interrogation were assessed every 6 months, overall and stratified by in-person versus remote management. Rhythm monitoring and post-implant follow-up were determined based on billing codes (**Supplemental Table 2**).

### Statistical Analysis

We calculated annual incidence of ILR placement per 100,000 at-risk patient-days, stratified by indication and inpatient vs. outpatient setting. We assessed temporal trends in ILR placement by estimating the relative annual change in incidence. Mean relative annual change was estimated using Poisson regression with a log transformed offset term that accounted for the population at risk. Regressions were performed for all ILRs, by guidance concordance category and by clinical indication, calculating estimates with 95% confidence intervals.

We calculated descriptive statistics of all patients receiving ILR based on age, sex, ethnicity, race, payor, social vulnerability index (SVI), and urbanicity. Urbanicity and SVI were both assigned based on patient zip code. Urbanicity was defined by rural-urban commuting area (RUCA) codes, classified as metropolitan (RUCA<4), micropolitan (RUCA 4-6), small town (RUCA 7-9), and rural (RUCA 10). We queried Cosmos data using SQL and performed analyses in R version 4.5.1. The study was deemed exempt by the Yale IRB (Category 4). Epic Cosmos contains fully de-identified EHR data, and therefore no identifiable private information was accessed.

## Results

### Population-level trends in ILR placement, overall and by setting and guidance concordance

There were 47,046,706 patients across 142 unique health systems with any diagnosis for a clinical indication within one of the 13 potentially eligible indication categories for ILR placement, of whom 201,103 (0.43%) underwent ILR placement between January 1, 2020 and December 31, 2025 (**Figure 1**). These patients accounted for 98.6% of all ILR placements (total of 203,866) over this period, as 2,763 had not received any of the 13 potentially eligible diagnoses. The annual incidence in 2020 was 3.08 ILR placements per 100,000 patient-days, compared to 4.13 ILRs per 100,000 patient-days in 2025, a relative annual increase of 5.9% (95% CI, 5.6%-6.1%; p<0.001).

**Figure 1:**
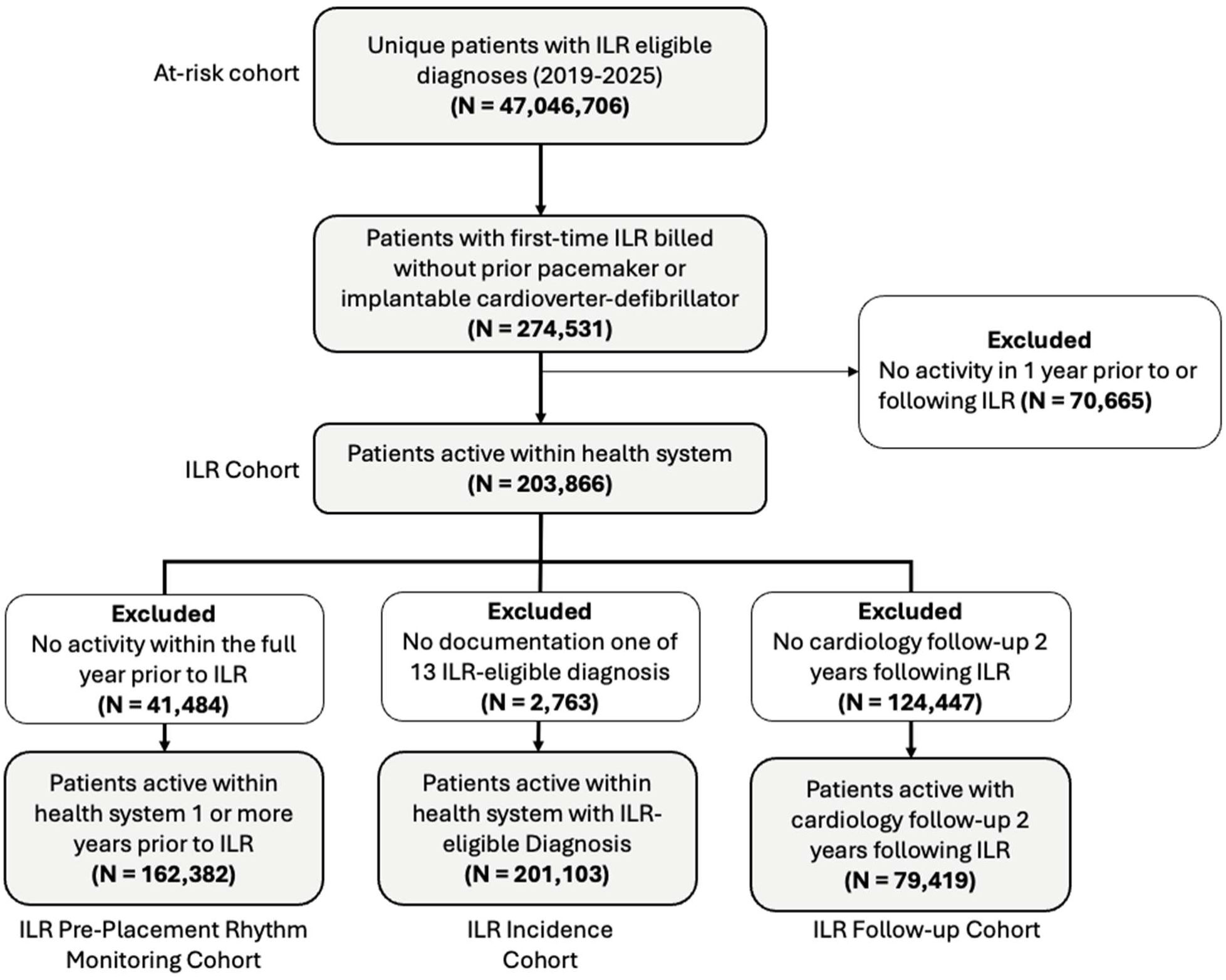
Flowchart of Patient Cohort. Abbreviations: Implantable loop recorder (ILR)

Approximately three-fourths of patients with an eligible diagnosis, 154,892 (77.0%), underwent ILR placement in outpatient settings. Annual incidence of outpatient ILR placements increased over the study period, from 2.32 ILR placements per 100,000 patient-days in 2020 to 3.20 in 2025, a relative annual increase of 6.2% (95% CI, 5.8%-6.5%; p<0.001) **(Figure 2 and Supplemental Table 4)**. Annual incidence of inpatient ILR placements increased more modestly over time, from 0.76 ILR placements per 100,000 patient-days in 2020 to 0.93 in 2025, a relative annual increase of 5.0% (95% CI, 4.4%-5.5%; p<0.001).

**Figure 2:**
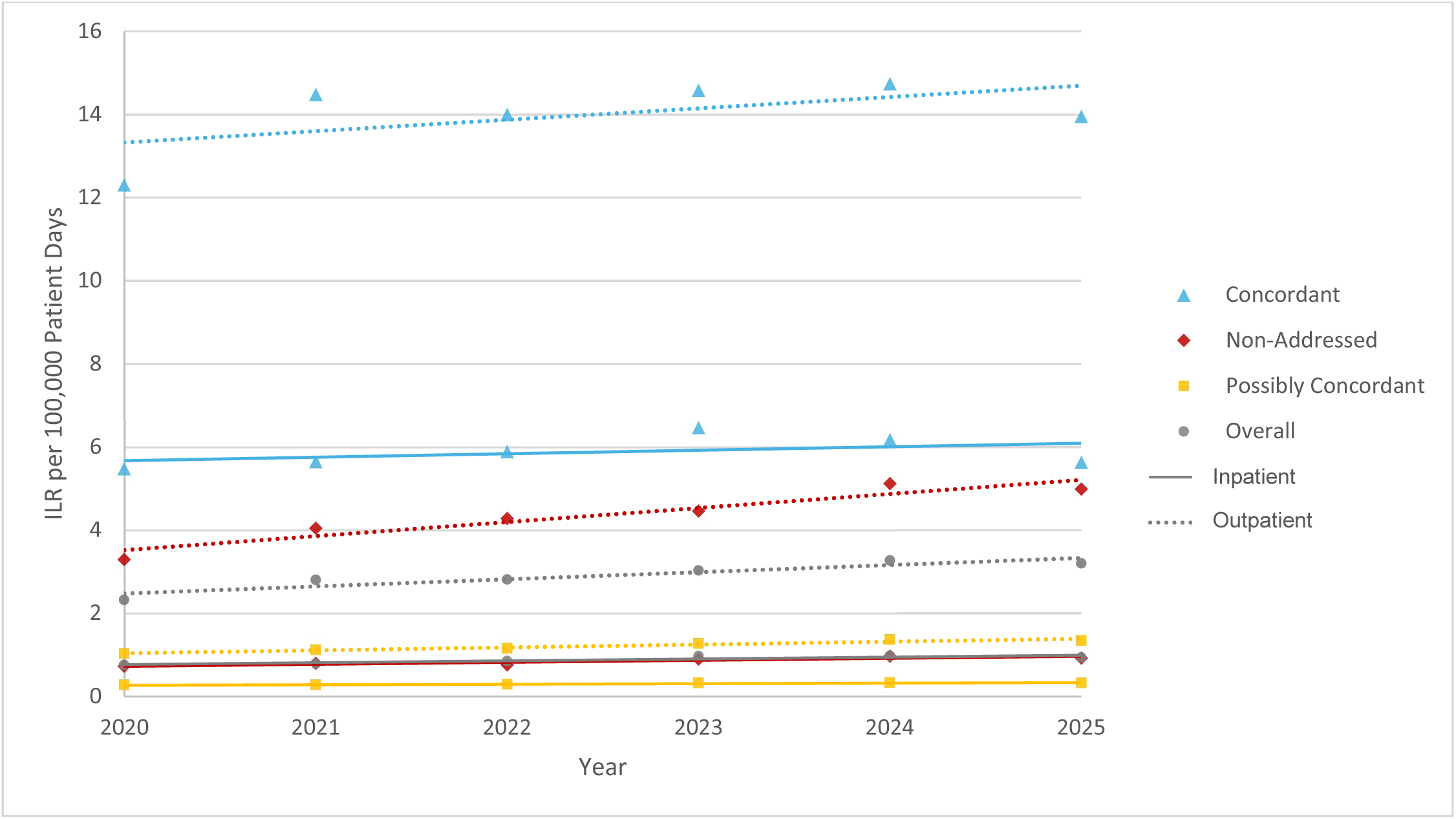
Incidence of Implantable Loop Recorder Placement by Guidance Concordance Classification, Stratified by Inpatient vs Outpatient Setting, 2020 to 2025.

There was variation in annual incidence of ILR placements by clinical indication, ranging from 0.04 ILRs per 100,000 patient-days for patients with a diagnosis of recurrent falls to 27.21 for patients with a diagnosis of cryptogenic stroke in 2025 (**Table 2**).

**Table 2.** Implantable Loop Recorder Placement Incidence and Change, Stratified by Indication and Guidance Concordance Across All Clinical Settings.

| **Guidance-Aligned Grouping** | **2020 Incidence**<br>(ILR per 100,000 patient-days) | **2025 Incidence**<br>(ILR per 100,000 patient-days) | **Relative Annual Change, 2020-25 (95% CI)** |
| --- | --- | --- | --- |
| *All Diagnoses* | **3.08** | **4.13** | **5.9%**<br>**(5.6, 6.1)** |
| *Guidance Concordant* | **17.80** | **19.59** | **1.3%**<br>**(0.9, 1.7)** |
| Recurrent syncope | 13.85 | 12.99 | -1.8%<br>(-2.5, -1.1) |
| Cryptogenic stroke | 20.73 | 27.21 | 5.1%<br>(4.5, 5.5) |
| *Possibly Guidance Concordant* | **1.30** | **1.67** | **5.5%**<br>**(5.0, 6.0)** |
| AF surveillance (within 90 days post-ablation) | 13.48 | 10.60 | -3.6%<br>(-5.1, -2.1) |
| Stroke or TIA | 3.45 | 3.77 | 1.7%<br>(0.7, 2.8) |
| Syncope | 2.11 | 2.06 | -0.3%<br>(-1.2, 0.7) |
| Non-sustained VT or frequent PVC | 1.95 | 3.19 | 11.2%<br>(9.9, 12.4) |
| Palpitations | 1.28 | 1.38 | 2.3%<br>(1.2, 3.5) |
| Uncontrolled epilepsy | 0.27 | 0.39 | 7.1%<br>(3.1, 11.4) |
| Recurrent falls | 0.06 | 0.04 | -0.2%<br>(-9.4, 9.8) |
| Dizziness | 0.08 | 0.17 | 14.8%<br>(10.7, 19.1) |
| *Guidance Non-addressed* | **4.02** | **5.91** | **7.4%**<br>**(6.8, 8.0)** |
| AF monitoring (no ablation) | 5.43 | 8.86 | 10.0%<br>(9.4, 10.7) |
| Atrial flutter | 2.06 | 2.90 | 5.3%<br>(3.3, 7.4) |
| SVT | 0.97 | 1.50 | 7.2%<br>(4.9, 9.4) |

Atrial fibrillation (AF), Implantable loop recorder (ILR), Premature ventricular contraction (PVC), Supraventricular tachycardia (SVT), Transient ischemic attack (TIA), Ventricular tachycardia (VT),

Annual incidence of ILR placement for clinical indications classified as guidance concordant increased significantly over time at the lowest relative rate, from 17.80 ILR placements per 100,000 patient-days in 2020 to 19.59 in 2025, a relative annual increase of 1.3% (95% CI, 0.9%-1.7%; p<0.001). ILR placement for clinical indications classified as possibly guidance concordant increased significantly over time at a moderate relative rate, from 1.30 ILRs placed per 100,000 patient days in 2020 to 1.67 in 2025, a relative annual increase of 5.5% (95% CI, 5.0%-6.0%; p<0.001). Annual incidence of ILR placement for clinical indications classified as guidance non-addressed increased significantly over time at the highest relative rate, from 4.02 ILRs per 100,000 patient days in 2020 to 5.91 in 2025, a relative annual increase of 7.4% (95% CI, 6.8%-8.0%; p<0.001).

### Characteristics of Patients who Underwent ILR Placement

Among the 203,866 patients who underwent ILR placement for any indication, mean age was 67.7 (standard deviation [SD] 13.7) years; 97,389 (47.8%) were male; 141,781 (69.5%) were White, 19,091 (9.4%) were Black, and 27,351 (13.4%) were another race or multiple races; and 39,564 (19.4%) patients were in the most socially vulnerable quartile **(Table 3)**. The primary payor was Medicare for 74,049 (36.3%), Medicare Advantage for 51,245 (25.1%), other commercial payor for 49,162 (24.1%), and Medicaid for 15,293 (7.5%).

**Table 3.** Demographic Characteristics of Patients Receiving Implantable Loop Recorders.

| **ILR placement (N = 203,866)** |  |
| --- | --- |
| **Mean age at first ILR placement (years)** | 67.7 (SD = 13.7) |
| <45 | 14,362 (7.0%) |
| 45-54 | 17,830 (8.7%) |
| 55-64 | 36,074 (17.7%) |
| 65-74 | 64,528 (31.7%) |
| >75 | 71,072 (34.9%) |
| **Sex** |  |
| Male | 97,389 (47.8%) |
| Female | 104,941 (51.5%) |
| Other | 1,536 (0.8%) |
| **Ethnicity** |  |
| Hispanic | 8,731 (4.3%) |
| Non-Hispanic | 174,916 (85.8%) |
| Other Ethnicity | 20,219 (9.9%) |
| **Race** |  |
| American Indian or Alaskan | 574 (0.3%) |
| Asian or Pacific Islander | 2,178 (1.1%) |
| Black | 19,091 (9.4%) |
| Other Single Race* | 3,408 (1.7%) |
| White | 141,781 (69.5%) |
| Multiple Races | 21,191 (10.4%) |
| Missing | 15,643 (7.7%) |
| **Payor Type** |  |
| Traditional Medicare | 74,049 (36.3%) |
| Medicare Advantage | 51,245 (25.1%) |
| Commercial | 49,162 (24.1%) |
| Medicaid | 15,293 (7.5%) |
| VA / TRICARE | 3,273 (1.6%) |
| Other Payor | 9,520 (4.7%) |
| Missing | 1,324 (0.6%) |
| **Social Vulnerability Index (SVI) percentile ranking** |  |
| 1 (most vulnerable) | 39,564 (19.4%) |
| 2 | 48,067 (23.6%) |
| 3 | 50,698 (24.9%) |
| 4 (least vulnerable) | 61,244 (30.0%) |
| Missing | 4,293 (2.1%) |
| **Urbanicity by Rural-Urban Commuting Area (RUCA)** |  |
| Metropolitan areas (RUCA < 4) | 164,726 (80.8%) |
| Micropolitan (4-6) | 18,328 (9.0%) |
| Small towns (7-9) | 10,064 (4.9%) |
| Rural (10) | 6,710 (3.3%) |
| Missing | 4,038 (2.0%) |

Notes: SVI and RUCA scores assigned within Epic Cosmos based on patient zipcode; “missing” denotes data not available in Epic Cosmos, while “other” indicates existing data that did not fit into demographic categories listed in the table above.

### ILR Placement by Clinical Indication

Among the 203,866 patients who underwent ILR placement during the study period, 89,462 (43.9%) underwent ILR placement for clinical indications classified as guidance concordant, 58,060 (28.5%) possibly concordant, 50,541 (24.8%) guidance non-addressed, and 5,803 (2.8%) non-classifiable, among which 2,763 (1.4%) did not have any of 13 pre-identified diagnoses billed) **(Table 4)**.

**Table 4.** Placement Indications Among Patients Receiving Implantable Loop Recorders, 2020-2025.

| **Indication** | **Count** | **% of total (n = 203,866)** |
| --- | --- | --- |
| ***Definitely Guideline or Consensus Concordant*** | **89,462** | **(43.9%)** |
| Recurrent Syncope | 31,108 | (15.3%) |
| Cryptogenic Stroke | 58,354 | (28.6%) |
| ***Possibly Guideline or Consensus Concordant*** | **58,060** | **(28.5%)** |
| AF Surveillance (post-ablation) | 5,985 | (2.9%) |
| Stroke or TIA | 11,913 | (5.8%) |
| Syncope | 15,397 | (7.6%) |
| VT or PVC | 10,824 | (5.3%) |
| Palpitations | 11,847 | (5.8%) |
| Uncontrolled Epilepsy | 880 | (0.4%) |
| Recurrent Falls | 152 | (0.1%) |
| Dizziness | 1,062 | (0.5%) |
| ***Guideline or Consensus Non-addressed*** | **50,541** | **(24.8%)** |
| AF Monitoring (no-ablation) | 43,606 | (21.4%) |
| Atrial Flutter | 3,686 | (1.8%) |
| SVT or other Atrial Arrhythmia | 3,249 | (1.6%) |
| ***Non-classifiable*** | **5,803** | **(2.8%)** |
| Cryptogenic Stroke | 993 | (0.5%) |
| AF | 541 | (0.3%) |
| Stroke or TIA | 242 | (0.1%) |
| Syncope | 597 | (0.3%) |
| VT or PVC | 137 | (0.1%) |
| Palpitations | 359 | (0.2%) |
| Uncontrolled Epilepsy | 5 | (0.0%) |
| Recurrent Falls | 6 | (0.0%) |
| Dizziness | 38 | (0.0%) |
| Atrial Flutter | 44 | (0.0%) |
| SVT or other Atrial Arrhythmia | 78 | (0.0%) |
| Other uncategorized diagnosis billed | 2,763 | (1.4%) |

**Abbreviations:** Atrial Fibrillation (AF), Premature Ventricular Contraction (PVC), Supraventricular Tachycardia (SVT), Transient ischemic attack (TIA), Ventricular Tachycardia (VT)

The most common indication for ILR placement was cryptogenic stroke (n=58,354; 28.6%), a guidance concordant diagnosis. This was followed by atrial fibrillation (n=49,591; 24.3%); among which 43,606 were guidance non-addressed as they were not associated with an ablation within 90 days. Among possibly guidance concordant diagnoses, syncope (7.6%), stroke or TIA (5.8%), palpitations (5.8%), VT or PVC (5.3%), and AF surveillance within 90 days post-ablation (2.9%) were most common.

### Rhythm Monitoring Prior to ILR Placement

Among the 203,866 patients who underwent ILR placement, 162,382 (79.7%) had evidence of activity within the health system at least one year prior to ILR placement, 125,631 of which were outpatient placements. Among these 125,631 patients, 60,201 (47.9%) had documented prior rhythm monitoring. The longest monitoring duration was short-term (≤2 days) for 5,111 patients (4.1%), medium-term (>2 to ≤7 days) for 4,909 patients (3.9%), and longer-term monitoring (>7 days) for 50,181 patients (39.9%) **(Supplemental Figure 1)**. For patients with prior rhythm monitoring, median time from the evaluation of most recent monitoring to ILR placement was 86 (interquartile range: 47-159) days.

### ILR Follow-up

Among the 203,866 patients who underwent ILR placement, 79,419 (39.0%) had cardiology follow-up for at least 2 years following device placement, 15,436 (19.4%) of whom underwent ILR removal during that 2-year period. Of those 63,983 patients without ILR removal, 53,580 (83.7%) had any billed interrogation (55.6% remote only, 5.0% in-person only, 23.2% both) and 14,197 (22.2%) had any billed ILR reprogramming (0.3% remote only, 21.9% in-person only, <0.1% both) **(Supplemental Figure 2)**. In the first 6 months post-ILR placement, 75.7% had ≥1 interrogation (15.9% had 1–2 interrogations, 17.5% had 3–4, and 42.3% had ≥5), which declined to 57.1% in months 19–24 (14.4% had 1–2 interrogations, 13.5% had 3–4, and 29.2% had ≥5 during that 6-month period) **(Supplemental Figure 3)**.

## Discussion

In this retrospective cohort study using electronic health record data from more than 140 health systems from 2020–2025, we found that ILR placements were uncommon, as fewer than 1% of all patients with diagnoses potentially eligible for ILR placement received an ILR. Nevertheless, ILR placements have increased steadily over time, with larger annual growth for patient indications that were guidance non-addressed, such as AF without recent ablation. These findings suggest that the growing use of ILR placements for clinical indications with lower strength evidence, as observed in prior research,^19^ has continued and highlights the importance of better research to understand the strengths and limitations of ILR placement for these clinical indications, particularly relative to noninvasive rhythm monitoring, where the evidence or professional guidance is less certain about their use. Additionally, fewer than half of patients had longer-term rhythm monitoring prior to outpatient ILR placement, suggesting that there may be an opportunity for more use of non-invasive rhythm monitoring, particularly for indications with less well-support.

Among indications not addressed by clinical guidelines or expert consensus statements, AF detection was the most common indication for ILR placements and had the fastest growth. More than one-fifth of ILRs were placed for AF monitoring without recent ablation. While ILRs provide long-term assessment of AF burden and duration to inform decisions regarding anticoagulation or antiarrhythmic drug use,^24,25^ there is not yet randomized clinical trial evidence that the more intensive monitoring provided by ILRs compared with non-invasive approaches improves clinical outcomes.

Clinical guidance recommends a step-wise approach for rhythm monitoring in multiple indications, with use of non-invasive cardiac monitoring before considering ILR.^10–12,14,26^ In contrast, our study suggests that ILRs are often employed as first-line rhythm monitoring modalities for slightly more than half of outpatients. One possible explanation for the relatively low rates of rhythm monitoring is that ILR placement was preceded by findings on wearable devices, which are increasingly adopted and have regulatory clearance for identifying AF.^20,27,28^

The smaller size, ease of placement, and AI algorithms for ILRs^1,3,6,25,29^ should also be considered alongside rapid improvements in non-invasive monitoring technologies.^30–32^ These include recent data that patient-led smartwatch monitoring after AF ablation can reduce time to AF recurrence detection.^31^ The relative role of ILRs versus non-invasive rhythm monitoring, including wearable devices, should be more carefully compared for different indications to shape future rhythm monitoring strategies. These studies should also evaluate clinical burden and cost, given concerns about data review burden, false positives, and variable accuracy of these technologies.

We found that more than 80% of patients with ILRs had an ILR interrogation billed for within 2 years post-placement, and only three-fourths of ILRs were interrogated in the first 6 months despite being followed in cardiology care. These rates are lower than would be expected and may reflect real-world inconsistencies in device follow-up. Additionally, while recent ILR technology includes remote reprogramming capabilities designed to reduce false positives and transmission burden,^5^ use of remote programming was rare (0.3% of ILRs) despite its availability. These results differ from real-world analyses by ILR manufacturers that report similar overall programming rates but with most reprogramming occurring remotely.^4,5,33^ The discrepancy between our findings and those reports likely reflects infrequent or inconsistently used billing codes for remote programming rather than true absence of use.

Our study should be considered in the context of its limitations. First, the COVID pandemic affected care in the first years of the study, particularly 2020. However, by calculating incidence of ILR placement relative to diagnoses by patient setting, we control for changes in patient volume. Second, because we relied on electronic health record data, limited clinical details about ILR placement are available and coding errors are possible. Third, there may be incomplete information about prior rhythm monitoring and ILR follow-up because some patients receive care at other health systems not included in Epic Cosmos. We mitigated this issue by requiring sustained patient activity and assigning ILR indications only when diagnoses were coded twice in the outpatient setting; however, at-risk patients did not require a second diagnosis, making that group less precisely defined. It is possible that patients excluded for insufficient activity may differ systematically from those included. Fourth, each ILR placement was assigned a single primary diagnosis, although patients may have had multiple indications. To mitigate this issue, we coded the indication that was the most guidance concordant or of the greatest clinical severity. Fifth, classification of diagnoses for ILR placement relied on a 90-day window, which, while corresponding to prior studies,^19^ differs across patients and indications. However, this timeframe is likely reasonable because we would expect approximately 3 months to be sufficient for clinical decision-making in most circumstances.

## Conclusion

While ILR technology continues to advance, ILRs are increasingly placed for indications with lower-strength evidence, underscoring the need for continued evaluation of their comparative effectiveness relative to noninvasive rhythm monitoring.

## Data Availability

Epic Cosmos data were used for this analysis. These data originate from Epic electronic health records and cannot be shared directly by our study team. Access to Epic Cosmos data is available to researchers at institutions that participate in the Epic Cosmos network.

https://cosmos.epic.com/

## Acknowledgements

Matthew C. Andersen contributed to study conceptualization, all data analysis, visualization, original drafting of the manuscript, and review of cardiology guidelines. Sanket S. Dhruva, Joseph S. Ross, and Luis Correia provided research supervision, methodological guidance, and critical editing of the manuscript. Rohini Ghosh assisted with manuscript drafting and the review of cardiology guidelines.

